# Women’s experiences of emergency post-abortion care at Kawempe National Referral Hospital, Uganda: a qualitative descriptive study

**DOI:** 10.64898/2026.08.25.26360981

**Authors:** Kawungu Saad Sessimba, Aburi Godfrey James, Baluku Andrew, Igirimbabazi Pious, Kibuuka Balikuddembe, Keesiga Annette, Herbert Kayiga

## Abstract

**Background:** Post-abortion care (PAC) encompasses emergency treatment, counselling, contraceptive services, and referral linkages. Emergency post-abortion care (EPAC), the life-saving component of PAC, addresses acute abortion-related complications, including haemorrhage, sepsis, retained products of conception, and severe pain. In Uganda, where abortion is legally restricted and socially stigmatised, women’s care experiences are shaped by clinical urgency, fear, moral vulnerability, provider interactions, and structural health system constraints. Despite EPAC’s centrality to maternal survival, qualitative evidence on how women interpret and evaluate their care experiences in referral hospital settings in Uganda remains limited. This study explored women’s experiences of EPAC at Kawempe National Referral Hospital (KNRH) and identified the factors that shaped those experiences.

**Methods:** A qualitative descriptive design employing inductive thematic analysis was used. Sixteen in-depth interview transcripts from women who received EPAC at KNRH in March– April 2026 were analysed. The Socio-Ecological Model (SEM) was subsequently applied as a post-hoc organising framework to situate findings across individual, interpersonal, facility, and community levels.

**Results:** Six themes were identified: (1) survival and physical relief as the immediate measure of good care; (2) pain, fear, and emotional distress during treatment; (3) reassurance and support as buffers against vulnerability; (4) dignity under pressure: communication and privacy in EPAC; (5) structural barriers across the pathway of care; and (6) experiences beyond discharge: incomplete recovery and uncertainty. Care was frequently evaluated through the lens of survival, yet these accounts co-existed with intense procedural pain, compromised privacy, delays, financial burden, and inadequate post-discharge support. EPAC at KNRH was experienced as a complex encounter shaped by bodily vulnerability, interpersonal dynamics, and system-level constraints.

**Conclusions:** Strengthening EPAC requires patient-centred approaches that integrate clinical effectiveness with respectful communication, pain management, improved triage, and structured post-discharge support.

## Introduction

Post-abortion care (PAC) encompasses emergency treatment, counselling, contraceptive services, and referral linkages [1]. Emergency post-abortion care (EPAC), the clinical backbone of PAC, involves urgent management of women presenting with acute abortion-related complications such as haemorrhage, sepsis, retained products of conception, shock, or severe pain [2]. Unlike routine reproductive health services, EPAC is delivered in time-critical contexts where rapid stabilisation takes priority. Yet the quality of care is not determined by technical outcomes alone. Global evidence increasingly recognises that women evaluate care through a broader lens, including communication, dignity, privacy, emotional support, and timeliness [3, 4].

Globally, unsafe abortion accounts for a substantial proportion of maternal morbidity and mortality, predominantly in low- and middle-income countries. Globally, an estimated 45.1% of the 55.7 million abortions performed annually between 2010 and 2014 were unsafe, defined by WHO as a procedure to terminate a pregnancy carried out either by a person lacking the necessary skills or in an environment not meeting minimal medical standards, or both, generating complications that require emergency medical intervention [1, 5]. In these settings, EPAC serves as a vital entry point into the health system, and timely access is strongly associated with reduced maternal morbidity and mortality. However, research consistently shows that survival does not guarantee a positive care experience: women frequently report fear, confusion, lack of information, and perceived mistreatment, even when clinical management is technically effective [6, 7].

In Sub-Saharan Africa (SSA), these challenges are compounded by health system constraints, including limited resources, high patient volumes, and workforce shortages. Women’s experiences of post-abortion care across the region are shaped not only by service availability but by the quality of delivery, particularly respectful communication, pain management, and privacy protections [8–10].

In Uganda, the context of EPAC is further shaped by restrictive abortion laws and strong social stigma surrounding abortion and pregnancy loss [11, 12]. Women may delay seeking care due to fear of judgment or legal consequences, and upon presentation, may carry feelings of guilt or shame. These emotional and social dimensions interact with the clinical encounter, shaping how care is experienced, remembered, and evaluated [5, 11].

Uganda’s legal restriction of abortion is often described narrowly in terms of criminal liability, but its consequences extend into the domains of the right to health, bodily self-determination, and timely access to emergency care. Bioethical and medico-legal scholarship increasingly frames restrictive abortion regimes as a determinant of health inequity rather than solely a matter of criminal law [13]. Empirically, a systematic review of abortion law reforms across low- and middle-income countries found that liberalisation was generally associated with improved health-service access and outcomes, while restrictive regimes were linked to delayed care-seeking and greater reliance on unsafe methods [14]. Read together, this evidence positions Uganda’s restrictive legal environment not merely as a backdrop to individual women’s fear and stigma, but as a structural determinant that shapes whether, when, and how safely women reach EPAC in the first place.

KNRH, one of Uganda’s busiest public maternity facilities, receives a high volume of women with abortion-related emergencies, handling over 200 EPAC cases monthly [15]. Women frequently arrive after having navigated multiple lower-level facilities that lack essential resources such as blood, equipment, or trained personnel [16]. Evidence from related obstetric contexts at KNRH shows that even when clinical care is effective, women may still experience poor communication and emotional neglect [17].

Furthermore, the EPAC experience extends beyond discharge. Women may continue to experience physical symptoms, uncertainty about recovery, concerns about future fertility, and varying levels of social support or stigma within their communities. Evidence from related post-discharge and postnatal care contexts suggests that care experiences influence not only immediate recovery but also future health-seeking behaviour and trust in the health system [18, 19].

Despite EPAC’s central role in maternal health, most research in Uganda has focused on clinical outcomes, service availability, and utilisation patterns [20–22]. Without evidence on how women themselves interpret and evaluate their care experiences, particularly in emergency contexts, quality improvement efforts risk prioritising technical performance while overlooking the relational, emotional, and structural factors that international quality-of-care frameworks increasingly recognise as central to patient experience [23].

Existing Ugandan evidence on post-abortion care is predominantly quantitative and utilisation-focused: national and sub-national studies have examined service availability, uptake, and the demographic correlates of utilisation [20–22], while the one qualitative account of patient-reported experience in a comparable tertiary setting focused on satisfaction rather than the interpretive, multi-level processes underlying it [23]. None of this literature addresses EPAC specifically as a time-critical emergency presentation, and none examines how women themselves interpret and weigh survival, dignity, and structural constraint against one another within a single national referral hospital. This study addresses that specific gap: a qualitative, multi-level account of how women experience EPAC as an emergency encounter, rather than a satisfaction or utilisation outcome.

This study, therefore, explored women’s experiences of EPAC at KNRH through a qualitative lens, applying the Socio-Ecological Model (SEM) to examine factors operating at the individual, interpersonal, facility, and community levels.

## Materials and methods

### Ethics statement

This study was approved by the Makerere University School of Medicine Research and Ethics Committee (SOMREC; approval reference: Mak-SOMREC-2025-913) and by Kawempe National Referral Hospital. All participants provided written informed consent before the interview. Given the legally restricted status of abortion in Uganda, consent procedures included participants were explicitly informed, as part of consent, of the legal implications of audio-recorded disclosures; audio recordings were encrypted and stored separately from identifying information; and a ten-year retention period was specified, after which recordings will be deleted in line with the approved SOMREC protocol. Participants were assigned anonymised codes; no identifying information was included in transcripts or analysis. Participation was voluntary, participants were informed that declining or withdrawing would not affect their clinical care, and could withdraw at any time without consequence. Interviewers were trained to check with participants throughout the interview whether they wished to continue, and participants were permitted to pause or discontinue the interview at any point where distress made this necessary; this protocol was applied during data collection.

### Study design

A qualitative descriptive design was employed, using inductive thematic analysis to generate findings grounded closely in participants’ own accounts, without commitment to a specific phenomenological, grounded-theory, or narrative analytic tradition [24]. This design was considered appropriate for a first qualitative account of EPAC experiences at KNRH, where the priority was to represent women’s reported experiences in accessible, data-near terms. This approach enabled exploration of the physical, emotional, and social dimensions of women’s EPAC experiences, capturing depth and context not accessible through quantitative methods alone. Following inductive theme generation, the Socio-Ecological Model (SEM) was applied post hoc as an organising interpretive lens rather than as an independent analytic method or a deductive coding framework; the rationale for this combination is set out in Data analysis, below.

### Researcher positionality

All authors are clinician-researchers affiliated with the Department of Obstetrics and Gynaecology at Makerere University College of Health Sciences and/or Kawempe National Referral Hospital, the study site. This insider position provided contextual and clinical familiarity with EPAC service delivery, but also created the potential for professional norms regarding acceptable procedural pain, standard waiting times, and routine communication practices to shape which participant accounts were interpreted as analytically significant, and for institutional affiliation to influence how provider-level versus system-level explanations were weighted during interpretation. The interviews were conducted by the corresponding author, a postgraduate trainee in the same department, with research assistants who were registered midwives employed at KNRH facilitating participant approach and consent. No participant had a prior relationship with the interviewer or research assistants before the study: first contact for study purposes occurred at discharge, as part of the two-stage recruitment process described above, not through any pre-existing clinical relationship. Before consent was obtained, participants were told the interviewer’s role as a postgraduate researcher, the purpose of the study, and that participation was voluntary and would not affect their clinical care. Because both the interviewing team and the analytic team were clinically and institutionally affiliated with the service under study, this insider position extended to the interview encounter itself, not only to analysis: it created potential for professional norms regarding acceptable procedural pain, standard waiting times, and routine communication practices to shape what participants felt able to disclose to a clinically affiliated interviewer, as well as which accounts were subsequently interpreted as analytically significant. Formal coding and theme development were conducted by the corresponding author, with review by two senior co-authors. This review involved examining the coding log and analytic memo trail maintained throughout analysis by the corresponding author against the coded transcripts, and assessing theme–data fit, following initial theme development. Given that both the interviewing and analytic roles were held by clinician-researchers embedded in the study site’s own department, reflexivity was addressed through written reflexive journals and structured debriefing at three points in the research process. Before data collection began, the research team held a session in which each member recorded their own professional assumptions, clinical background, and expectations regarding EPAC care. During data collection, each interviewer (the corresponding author and the research assistants) kept a separate reflexive journal, writing a short entry after every interview covering the interview setting, the participant’s emotional state, and the interviewer’s own reactions and any surprises; because journals were kept individually rather than jointly, they captured differing positions across the team, including clinical and non-clinical, and senior and junior perspectives. During analysis, analytic memos were attached to transcripts and reviewed in team coding meetings, where interpretive disagreements were discussed and resolved. This process was used specifically to bracket clinical assumptions: because the corresponding author and other team members bring clinical health-systems backgrounds, assumptions likely to shape coding, for example what counts as “normal” procedural pain or an “acceptable” waiting time, were recorded before coding began, and codes were checked against participants’ own words rather than against clinical standards; where a code reflected the team’s own judgement, for example labelling an account as describing a “delay”, the underlying transcript was checked for whether the participant herself had framed the experience that way.

### Study setting

The study was conducted at KNRH in Kampala, Uganda, within the gynaecological emergency unit, where women with abortion-related complications are assessed, managed, and reviewed. KNRH is Uganda’s national referral centre for obstetric and gynaecological emergencies, handling over 200 EPAC cases per month (approximately seven cases per day) [15]. EPAC is provided free of charge, with out-of-pocket payments required only for ultrasound scans and medications that are out of stock. Services are available 24 hours a day, 365 days a year, delivered by a multi-cadre team ranging from senior consultants to student health workers. During the study period, privacy provisions within the unit were variable: participant accounts (see Theme 4, below) describe curtains being used during some examinations and procedures but not others, consistent with a shared clinical space rather than fully private cubicles for every patient. This description is derived from participant accounts rather than facility records.

### Study population and data source

This analysis draws on sixteen in-depth interview transcripts collected as part of a broader parent study of EPAC experiences at KNRH; the present study constitutes a secondary thematic analysis of these transcripts, conducted with the aims and analytic approach described in this Methods section. Participants ranged in age from 18 to 45 years and represented diverse socio-demographic and reproductive backgrounds, including variation in marital status, educational attainment, occupation, and parity. The term post-abortion care, as used in this study, encompasses presentations following both spontaneous pregnancy loss (miscarriage) and induced abortion; the clinical intake and interview protocol did not systematically record which of these applied for each participant, and this distinction was not used as an inclusion or exclusion criterion; this was genuinely not captured in the clinical or interview records available for this analysis. Consequently, interpretations concerning stigma, guilt, and moral vulnerability in the Results and Discussion are drawn only from narratives in which participants themselves disclosed or implied the nature of the pregnancy loss, and should not be read as characterising the full sample; this constraint is discussed further in Strengths and limitations, below.

### Sampling strategy

Purposive sampling with maximal variation was used to select participants who had direct experience of EPAC and were approached after at least 14 days had elapsed since discharge. This window allowed sufficient time for initial physical recovery and reflection while minimising recall bias, without approaching women during the acute treatment period. Eligibility required clinical stability at recruitment, informed consent, and willingness to participate in an interview conducted in English or Luganda. Maximal variation was operationalised using the demographic and clinical characteristics summarised in Table 1 (age, marital status, educational attainment, occupation, and parity); geographic origin relative to KNRH and religious affiliation were not used as sampling variables and are acknowledged as gaps in Strengths and limitations, below. A total of twenty-four women meeting eligibility criteria were approached during the study period, of whom sixteen consented and eight declined (see Data collection, below); this figure reflects women identified and approached within the two-month recruitment window rather than a further sub-sample drawn from a larger caseload.

**Table 1.**
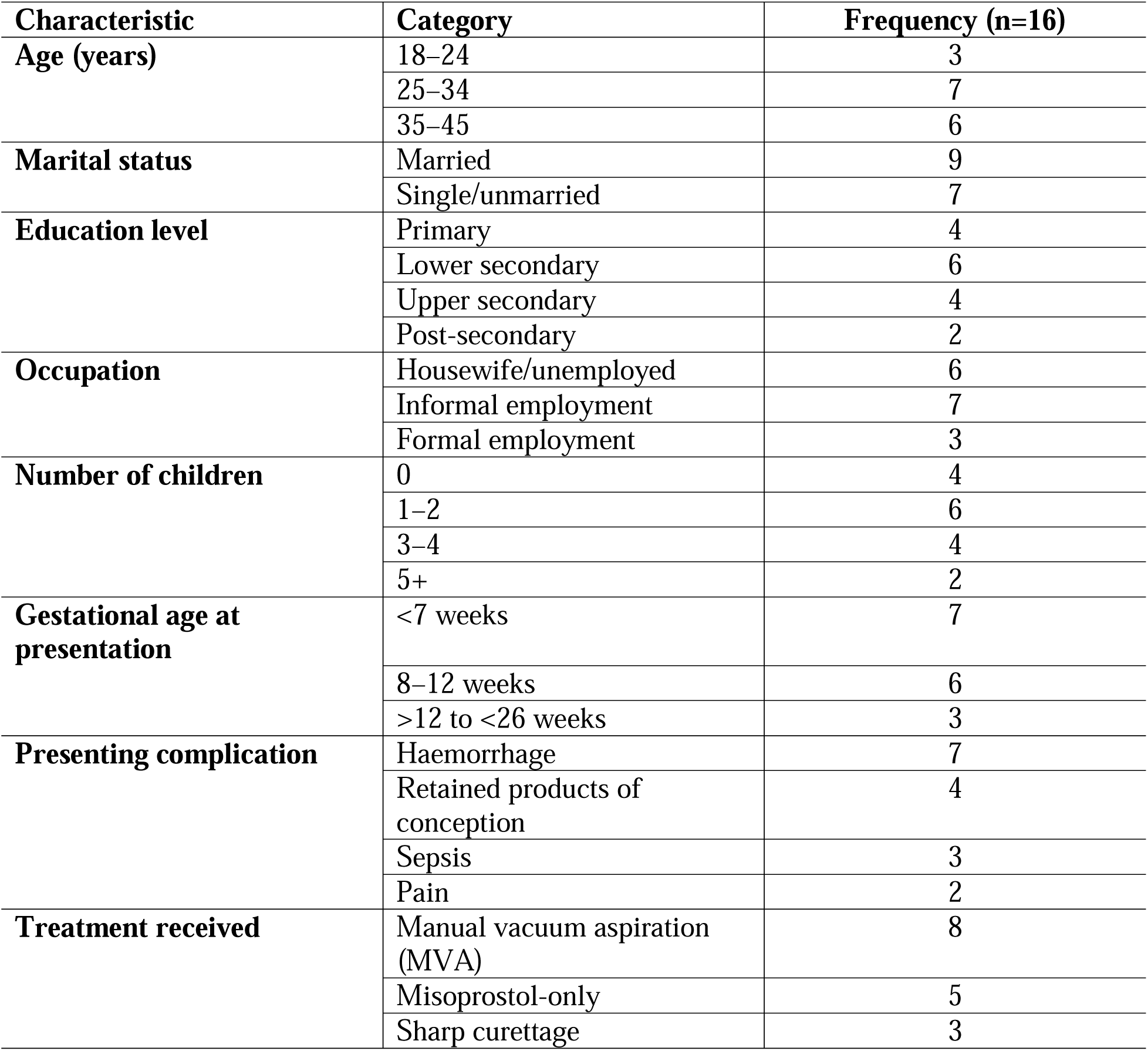
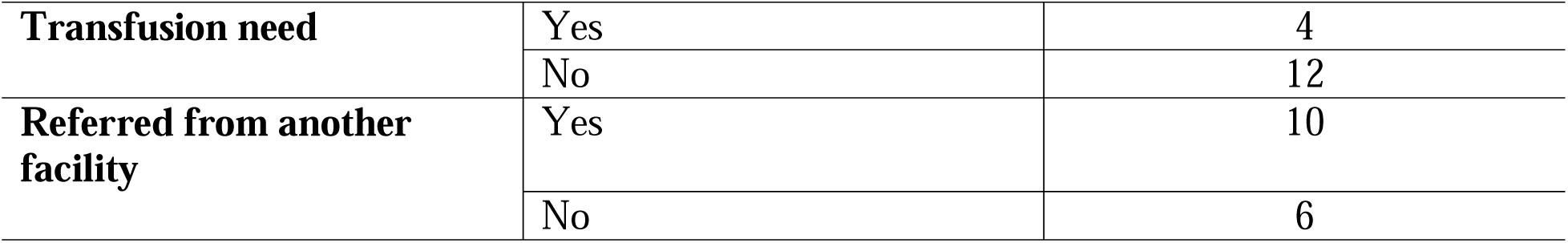
Socio-demographic characteristics of study participants (March–April 2026, n=16).

**Table 2.** Overview of themes, sub-themes, and meaningful units/codes.

| Theme | Sub-theme | Meaningful units (codes) |
| --- | --- | --- |
| **1. Survival and physical relief as the immediate measure of good care** | Physical stabilisation and symptom relief | Bleeding stopped; pain reduced; felt saved |
|  | Rapid emergency response | Attended quickly; prioritised; emergency action taken |
|  | Survival as minimum threshold | Gratitude for survival; care judged by outcome |
| **2. Pain, fear, and emotional distress during treatment** | Procedural pain and bodily suffering | Painful evacuation; severe discomfort; helplessness |
|  | Fear of death and acute uncertainty | Fear of dying; shock; severe bleeding; anxiety |
|  | Emotional conflict and self-judgment | Guilt; regret; self-blame; internal distress |
| **3. Reassurance and support as buffers against vulnerability** | Provider reassurance and emotional calming | Comfort; counselling; encouragement; calming words |
|  | Family and social support | Transport; money; caregiving; relative presence |
|  | Absence of support | Alone; abandonment; dependence; emotional isolation |
| **4. Dignity under pressure: communication and privacy in EPAC** | Respectful communication and explanation | Welcomed; informed; listened to; spoken to well |
|  | Dismissive or judgmental communication | Rude language; blame; harsh tone; not listened to |
|  | Privacy preserved or violated. | Curtain used; exposure; embarrassment; many observers |
| **5. Structural barriers across the pathway of care** | Delays and waiting under emergency conditions | Long waits; delayed review; bleeding while waiting |
|  | Overcrowding and limited resources | Many patients; lack of blood; staff shortage |
|  | Financial burden within care | Paying for scan; buying medicines; borrowing money. |
|  | Referral and transfer challenges | Moved between facilities; no blood; delayed care |
| **6. Experiences beyond discharge: incomplete recovery and uncertainty** | Persistent symptoms after discharge | Ongoing pain; bleeding; weakness; possible infection |
|  | Limited follow-up and continuity gaps | No review; no follow-up call; no check-up |
|  | Fertility and reproductive uncertainty | Future pregnancy concerns; uncertainty about safe conception timing |
|  | Social support, silence, and stigma after discharge | Selective disclosure; fear of judgment |

Sampling was guided by the principle of data saturation: data collection continued until no new codes, categories, or themes emerged from additional transcripts [25]. Saturation was assessed narratively: codes became consistent and repetitive across the later transcripts, and no new codes, categories, or themes emerged from the final transcripts reviewed. This narrative assessment follows the general saturation principle described by Guest, Namey, and Chen [23], but does not apply that paper’s quantitative tracking metrics (e.g., base size, run length, or new-code ratio); readers should treat the saturation claim as a qualitative judgement rather than a metrics-based determination. As this study used inductive thematic analysis rather than a phenomenological design, sample adequacy is assessed by thematic saturation rather than by phenomenological criteria such as depth of individual accounts.

### Data collection

Data were collected through semi-structured in-depth interviews following a two-stage recruitment process. In the first stage, women who had received EPAC were identified at discharge; their contact details were obtained, and a scheduled review appointment was confirmed. Recruitment deliberately targeted this scheduled review appointment, which in standard post-abortion clinical practice at the unit falls around two weeks after discharge, rather than the point of discharge itself, when women are often physically weak, emotionally distressed, and under time pressure to leave the facility; this is consistent with the interval between discharge and interview reported in Table 1, where all sixteen participants were interviewed at or beyond the 14-day mark. In the second stage, on or around the review date, the research team confirmed participants’ arrival at the hospital. Women who met eligibility criteria were approached by a trained research assistant, given a verbal explanation of the study, and offered time to consider participation before written informed consent was obtained. Interviews were conducted by the corresponding author, a postgraduate trainee in the same department, with research assistants who were registered midwives employed at KNRH facilitating participant approach and consent; researcher positionality is addressed below.

Of the women approached during the study period (March–April 2026), eight declined to participate citing time constraints or personal concerns; this 33% refusal rate is not further characterised here and is addressed as a potential source of selection bias in Strengths and limitations, below. The sixteen who consented were interviewed individually in a consultation room at the gynaecological emergency unit, prioritising privacy. The interview guide (S1 File) opened with an account of the participant’s overall experience of care, from first realising something was wrong until leaving the hospital, including probes on privacy, confidentiality, and feeling respected, and continued with: prior knowledge and expectations of care; interactions with health workers; communication and information-sharing; support from family and others; community attitudes and stigma; recommendations for the health system and policy; and a final open reflection. Interviews were conducted in English or Luganda according to participant preference, audio-recorded, and transcribed verbatim. Translation and back-translation of Luganda-language interviews were conducted by the interviewers themselves rather than by independent professional translators; this is noted as a limitation in Strengths and limitations, below.

### Data analysis

Transcripts were read repeatedly to develop familiarity with the data. Meaningful units of text were identified, coded, and grouped into categories based on similarities and patterns across accounts. Themes were generated inductively through thematic analysis, moving iteratively between participants’ language and broader interpretive concepts. Constant comparison across transcripts was used to identify both common patterns and divergent experiences [25].

The SEM was applied as an interpretive lens at later stages of analysis to organise findings by the level at which they operated: individual, interpersonal, facility, and community. This framework was originally developed to explain how human experiences are shaped across multiple nested levels of influence [26] and has since been applied to health service research to guide multi-level interpretive frameworks [27]. This approach, of using the SEM as an organising framework rather than a deductive coding template, allowed themes to remain grounded in participants’ narratives while being situated within a theoretically coherent multi-level account.

Combining inductive thematic analysis with a subsequently applied SEM organising lens follows established framework-application approaches in applied qualitative health research, in which a theoretical model is used to structure the presentation of inductively derived findings rather than to generate them. Each theme was assigned to the SEM level at which its central content was most directly located. For example, bodily and psychological experience at the individual level, provider–patient communication at the interpersonal level, resource and service-delivery conditions at the facility level, and stigma and social norms at the community level. Themes with content spanning more than one level (notably Theme 6) were retained under their most proximate level, with cross-level connections addressed narratively in the Discussion rather than through re-coding. The SEM is therefore presented as an organising interpretation of the thematic data rather than as an independent source of findings, and the Discussion distinguishes between conclusions grounded directly in participants’ accounts and those introduced through the SEM’s organising structure.

### Rigour and trustworthiness

Rigour was ensured by applying Lincoln and Guba’s criteria for trustworthiness: credibility, dependability, and transferability [24, 28]. Credibility was enhanced through immersive, iterative engagement with participants’ own words, supplemented by senior-author review of theme– quote correspondence, which functioned as informal peer debriefing. Dependability was supported through consistent application of the coding framework, and through a coding log and analytic memo trail maintained throughout analysis by the corresponding author, available to reviewers on reasonable request. Confirmability, the fourth criterion in Lincoln and Guba’s framework, was addressed through the same coding log and memo trail, which document the progression from raw data to final themes and can be made available to reviewers to support independent audit. Transferability was supported by detailed descriptions of the study setting, participant characteristics, and analytical process, bounded by the single-site, single-facility nature of the study (see Strengths and limitations, below). Transparency was maintained by attending to both positive and negative experiences, including contradictions within individual accounts, to avoid selective reporting. The single-analyst coding conducted by the corresponding author was reviewed by two senior co-authors, involving independent review of the coding log and memo trail against the coded transcripts to assess theme–data fit, conducted following initial theme development; this served as the primary safeguard against single-analyst bias in this study.

## Results

### Participant characteristics

Sixteen in-depth interview transcripts were included in the final analysis. Participants ranged in age from 18 to 45 years and represented diverse social and reproductive backgrounds.

***Table 1*** presents socio-demographic characteristics.

### Overview of themes

All themes, sub-themes and corresponding meaningful units/codes are summarised in

### Theme 1: Survival and physical relief as the immediate measure of good care

For most women, the first and most immediate meaning of EPAC was survival. Participants described care as “good” when it produced recognisable bodily improvement such as relief from pain, reduced bleeding, or a sense that the condition had been controlled in time. Clinical recovery formed the earliest and strongest basis for evaluating care.

> *“They worked on me very well… they focused on me being fine. The doctors kept checking on me, and when I was told I could go home, I felt like a different person from the one who had come in. I was really happy with the way they treated me.” (IDI_16)*
>
> *“When I went to the hospital, I was feeling lower abdominal pain with every step I took, but after removing the pregnancy-related items from inside me, I felt relieved from the pain. It was like something heavy had been removed from inside me.” (IDI_6)*

The speed of response also shaped experience. Visible urgency from providers conveyed both seriousness and competence, reinforcing women’s confidence that care was effective.

> *“When I reached there, they all rushed, wanting to know what had happened to me. One doctor was already asking questions before I had even sat down, and another was already preparing to examine me. That reaction made me feel that my situation was being taken seriously.” (IDI_14)*

However, positive accounts were not uncomplicated. Women who described care as good still frequently narrated pain, fear, or emotional trauma within the same account, illustrating that survival, in the context of acute emergency, may temporarily overshadow shortcomings in communication, privacy, or emotional support.

### Theme 2: Pain, fear, and emotional distress during treatment

Despite the primacy of survival, women also described EPAC as intensely painful and emotionally distressing. Procedural pain emerged strongly, particularly during uterine evacuation.

> *“The moment when the machine was pulling the placenta out was so painful. It reached the point that when I heard metal being banged, I got scared and started shaking. I kept asking them to stop, but they told me to be patient because they were almost done.” (IDI_3)*
>
> *“The pain was too much when the doctor put it in the hand [inserted the intravenous cannula]; it was like my whole inside was being pulled out. I cried and asked him to stop, but he said he had to finish what he was doing. I sometimes dream of what he did to me, and I wake up feeling that same pain all over again.” (IDI_4)*

Fear of death was a recurring feature. Women who arrived haemorrhaging or acutely unwell often described uncertainty about what was happening within their bodies, intensifying fear and making treatment emotionally overwhelming.

> *“I thought I was going to die because of how much blood I had lost before reaching the hospital. But there was a female doctor who told me that people had come in worse conditions than mine and had survived. That calmed me a little, even though the fear was still there.” (IDI_16)*

Beyond physical suffering, some women described emotional conflict linked to the pregnancy event itself. Feelings of guilt, regret, or self-blame emerged across several transcripts, extending the meaning of distress beyond the procedure to include internal moral struggle.

> *“I didn’t like what I had done, and lying there in that bed made me think about it even more. I was feeling guilty and in pain at the same time, and there was nobody I could really talk to about it…” (IDI_16)*

### Theme 3: Reassurance and support as buffers against vulnerability

Support from healthcare providers, family, and social networks shaped women’s capacity to endure pain and fear. Provider reassurance emerged as a particularly important form of emotional support, functioning as a therapeutic intervention in its own right.

> *“The doctor took time and counselled me and comforted me, making me feel calm. He told me I would be fine, since people in worse conditions had survived and gone back to live their normal lives… I had arrived at the hospital believing I was going to die, and after he spoke to me, I started to think that maybe I would make it.” (IDI_2)*

Family support also played a major role, enabling women to access treatment, meet unexpected costs, and cope during recovery.

> *“My husband helped me throughout the whole time I was in the hospital. Whenever I needed a blood transfusion, he would provide the money, and if he didn’t have it, he would borrow from relatives or friends without complaining. Knowing he was there and willing to do whatever was needed gave me the strength to go through everything.” (IDI_2)*

Not all women had such support. Those who arrived alone described intensified vulnerability and complete dependence on staff.

> *“I had come alone because I did not want anyone to know why I was going to the hospital. But when I got there, and things became serious, I realised how much I needed someone. I couldn’t do anything on my own, and I had to depend completely on the nurses and doctors for everything. I was helpless…” (IDI_1)*

### Theme 4: Dignity under pressure — communication and privacy in EPAC

Women’s experiences were significantly influenced by whether they felt treated with dignity. Dignity was expressed through the quality of communication and the protection of physical privacy.

> *“He didn’t talk badly to me at all; he asked me what was wrong and even took me away from the crowded reception because I was in pain and began examining me immediately. When I asked him what he thought about my condition, he explained it until I understood, which I did not expect from a government hospital.” (IDI_11)*

However, other women experienced communication as dismissive or judgmental. In the context of abortion-related complications, such communication carried moral weight, deepening emotional discomfort.

> *“The doctor kept asking why I was alone and why we got pregnant at such a young age, and the way he was asking made me feel like I had done something very wrong. I wanted to explain myself, but he did not seem willing to listen. That made me feel ashamed on top of everything else I was already going through.” (IDI_1)*

Privacy was conditional: some women described curtains and restricted visibility; others reported feeling exposed during procedures.

> *“They worked on me in the open when everyone was seeing; there was no curtain and no screen between the other people in the room and me. I just felt embarrassed and kept looking away, hoping the people nearby were not watching… I had to endure it because I needed the treatment.” (IDI_1)*

### Theme 5: Structural barriers across the pathway of care

Women’s experiences were strongly shaped by structural conditions: delays, overcrowding, resource shortages, financial burden, and repeated movement across facilities before reaching definitive care.

> *“I waited for over 4 hours, and no doctor had checked on me, yet I was bleeding so much that I had to keep changing my clothes. Doctors were passing me by and not really listening when I tried to tell them how bad things were… We just sat there, I and two other women who had also been waiting a long time, all of us suffering in silence.” (IDI_10)*

Delays were not merely logistical; they became part of the emotional experience of illness, increasing fear and the feeling of neglect.

> *“If you don’t fight for your life, you can wait till evening without anyone bothering to check on you. I had to stand up twice and go and ask about my situation before they finally attended to me… not everyone has the strength to keep asking to be helped.” (IDI_12)*

Financial burden was a major issue. Women reported paying for scans and medications at a public facility, adding stress and uncertainty to emergency care.

> *“The doctor asked for money for the scan; it was sh.20,000, but I had only sh.15,000, and I didn’t know what to do. I was sick and worried, and now I had to think about money on top of everything else…” (IDI_6)*

These out-of-pocket costs take two distinct forms. The first is formal facility charges, for ultrasound scans and medications out of stock at the facility pharmacy, which are consistent with participants’ description of EPAC itself as officially free of charge. The second is informal payment: in at least one account (Theme 3, above), money was given directly to health workers not as a billed fee for the blood product or its screening, but as an incentive intended to motivate staff to act quickly in sourcing blood for an urgent transfusion. This distinction matters, because it indicates that even where EPAC and blood transfusion carry no official charge, women and their families may still experience a felt necessity to pay informally to secure timely emergency action, a pattern consistent with informal payment practices documented elsewhere in under-resourced public health systems. The women’s accounts indicate that this distinction between a nominally free procedure and its ancillary costs was not always apparent to them at the point of care, and itself became a source of distress.

Many women’s experiences began before arrival at KNRH, shaped by referral pathways involving multiple facilities. This content emerged through the interview guide’s open-ended opening question and its “what happened, step by step” probe, rather than through a dedicated question on referral; the guide contained no specific question on referral or transfer at any stage, whether before arrival or between departments within KNRH itself (e.g., between the emergency reception area, the gynaecological emergency unit, and theatre). The relative richness of pre-arrival referral content compared to the absence of intra-facility content may therefore reflect what participants spontaneously chose to narrate, rather than a designed protocol scope; the accounts reported here should accordingly be read as covering the pre-arrival referral pathway as it happened to be volunteered, not as a systematic account of referral experience. This gap in evidence on intra-facility movement is acknowledged as a limitation, below.

> *“They did not have blood to put on me at the first hospital, and yet I was losing so much that the doctor said I needed to get blood urgently. They told my husband to take me somewhere else… By the time I finally got the blood I needed, I had been moving from place to place for hours while getting weaker.” (IDI_2)*

### Theme 6: Experiences beyond discharge — incomplete recovery and uncertainty

Women’s experiences of EPAC did not end at discharge. Persistent physical symptoms, limited follow-up, unanswered questions, and variable social support shaped how they understood their recovery.

> *“Right now, I am feeling some pain, a little pain in my lower abdomen that has not gone away since I left the hospital. I also think I keep producing pus because there is something that comes out which does not look normal to me… nobody told me what to expect after going home.” (IDI_6)*

This account raised a possible symptom of infection disclosed during a research interview rather than a clinical encounter. The study protocol included a defined procedure for referring participants for clinical assessment where concerning symptoms were disclosed mid-interview; no case during data collection required this procedure to be invoked.

Follow-up care was largely absent, contributing to uncertainty about what was normal and when to seek help.

> *“No one has called me since I was discharged, not to check if I am okay or if the symptoms have changed. I thought maybe they would follow up in a day or two, but nothing came of it. After you’re discharged, that’s it.” (IDI_6)*

Uncertainty about future fertility and safe conception timing was a further source of distress that persisted after discharge, particularly where discharge counselling did not fully resolve women’s questions.

> *“My question was, if I want to conceive, what is the right period for me to get pregnant? They gave me a range, saying six months, eight months, but I wanted the exact answer because I do not want to make any mistakes. Not knowing the right time kept disturbing my mind even after I had left the hospital.” (IDI_15)*

Social context also mattered after discharge. Some women limited disclosure due to stigma, meaning that recovery was not only medical but also social and emotional.

> *“My mum called me, and I told her I was fine, because I did not want her to worry about the details of what had happened… I was not ready for what she would think of me…” (IDI_13)*

## Discussion

This study explored women’s experiences of EPAC at KNRH, Uganda, and identified the multi-level factors shaping those experiences. The findings reveal that women evaluated EPAC primarily through survival, with acute mortal threat leading women to prioritise survival as the primary evaluative criterion, temporarily subordinating relational and procedural quality dimensions. This pattern is consistent with earlier qualitative work [6, 7] and extends it by demonstrating that the primacy of survival in emergency settings was not merely a preference but a response to acute mortal threat. Importantly, this challenges the sole use of patient satisfaction surveys as quality indicators in EPAC, since women who survived life-threatening complications often rated care positively regardless of whether communication, pain management, or privacy were adequate [3, 4].

Once the immediate danger subsided, the embodied suffering that accompanied treatment came into sharper perspective. Pain in EPAC was not only a clinical symptom, but it was an interpretive experience carrying social, moral, and psychological weight. This convergence of physical and emotional suffering, documented by Coast et al. [29] and Ganatra et al. [5], was extended here into an emergency context, revealing that inadequate pain management amplified distress rather than containing it.

Reassurance from providers mediated between suffering and the capacity to endure it, functioning as a therapeutic intervention. When providers spoke calmly, explained what was happening, or remained emotionally present during painful procedures, women reported better ability to manage fear. This was consistent with Baum et al. [6, 30], Whitehouse et al. [31], and Chekol et al. [32]. In emergency settings, this function was amplified since women with severe haemorrhage were far more dependent on reassurance than those undergoing planned procedures. Absence of family support intensified this dependence [8, 18, 33].

In Uganda’s context of legal restriction and stigma, judgmental communication carried moral weight beyond poor service quality. It signalled condemnation to an already vulnerable woman. Cleeve [12] and Ouedraogo et al. [7] documented similar patterns; this study extended both findings by showing that in emergency settings where women cannot leave to seek care elsewhere, such communication functions as compounded disrespect, since women in acute emergency presentations cannot exit the care relationship, intensifying the impact of judgmental communication beyond contexts where patients retain the option to seek care elsewhere. Privacy violations compounded this by creating real social exposure risks in communities where abortion-related care is stigmatised.

Structural conditions constrained provider capacity for dignified care. Delays, shortages, and out-of-pocket costs at a public facility shaped the conditions under which relational care was possible, which was consistent with prior evidence [9, 16, 20]. Fragmented referral, a related pattern to the informal, self-sourced care-seeking pathways described by Ouma et al. [34], meant that for many women, EPAC had begun hours earlier under deteriorating conditions before reaching KNRH.

Discharge marked not the end of EPAC but a transition to largely unstructured recovery. Most prior Ugandan studies examined access and utilisation, not the post-discharge period [20–22]. Lythgoe et al. and Udho et al. [18, 19] showed that post-discharge experiences in SSA shaped future engagement with health services. This study confirmed and extended that finding specifically for EPAC: fertility-related uncertainty that persisted when discharge counselling failed to address conception timing constituted a psychological extension of the care experience itself.

Stillman et al. [35] examined post-abortion and safe-abortion care coverage, capacity, and caseloads during the global gag rule policy period in Ethiopia and Uganda, documenting service-level disruption during that period. Kabunga et al. [36] highlighted the added complexity for women living with HIV in northern Uganda, where fear, stigma, and moral conflict overlapped with the physical dimensions of care, extending the psychological meaning of EPAC well beyond the clinical encounter.

### Factors that influenced the women’s experiences guided by the SEM

Applying the SEM as an interpretive framework revealed that no single level fully accounted for women’s experiences; findings at each level were necessary but not sufficient.

At the individual level, bodily vulnerability, such as severity of haemorrhage, pain intensity, fear of death, and reproductive anxiety, shaped how women entered the care encounter and what they were able to perceive and evaluate. Women in extreme clinical distress were least able to assert preferences or notice deficiencies, and were most dependent on provider behaviour.

At the interpersonal level, provider communication, family support, and provider attitudes were decisive mediating factors, themselves shaped by facility-level conditions (overcrowding, staff pressures) and community-level factors (stigma). Cleeve [12] showed that provider stigma in Kampala shaped interactions with women seeking post-abortion care; this study extended that finding by tracing how stigma translated into specific communication practices that deepened women’s distress.

At the facility level, KNRH’s structural conditions, such as high patient volumes, blood product shortages, inadequate privacy arrangements, and absent follow-up systems, shaped the relational possibilities of care beyond its technical delivery, consistent with SSA-wide evidence [9, 20].

At the community level, abortion stigma in Uganda shaped the entire care trajectory: from delayed care-seeking, through morally loaded interpretations of provider behaviour, to post-discharge silence and selective disclosure. Ouma et al. [34] and Ndayisenga et al. [37] documented how stigma shapes abortion care pathways across East Africa; this study found that stigma operated within the facility during care and continued after discharge during recovery.

Beyond the community level, Uganda’s restrictive abortion legislation operates as a macro/policy-level determinant upstream of the community stigma, institutional risk aversion, and individual care-seeking delays documented across these findings, consistent with the Bronfenbrenner-rooted tradition underlying the SEM. This study did not directly elicit participant accounts at the policy level, so this connection is offered as an interpretive inference linking the findings to their broader legal context, rather than as a data-grounded finding in its own right.

The SEM provided a coherent multi-level account of the conditions under which clinical success and relational failure simultaneously occurred, making explicit the different systemic layers at which each operated: how women could simultaneously describe care as life-saving and inadequate. Survival was secured at the individual and facility levels through clinical intervention; dignity was undermined at the interpersonal level by poor communication and at the facility level by compromised privacy and delays; and community-level stigma amplified these indignities. The SEM demonstrated that EPAC quality was not a single outcome but a product of multiple, interacting, and sometimes contradictory forces operating simultaneously from crisis to post-discharge recovery.

### Strengths and limitations

This study is among the few qualitative investigations focused specifically on women’s experiences of EPAC at the national referral hospital level in Uganda, providing evidence directly relevant to policy and practice in a high-volume, high-acuity setting. The use of in-depth interview data and thematic analysis captured the depth, complexity, and contextual meaning of women’s experiences beyond what patient satisfaction surveys can access. Application of the SEM enabled a multi-level analysis that went beyond describing what women experienced to explaining the factors and interactions that produced those experiences. Inclusion of sixteen participants with diverse socio-demographic characteristics strengthened the analytical breadth of the findings.

The main limitation is that the study was conducted at a single site. All participants received care at KNRH, whose institutional characteristics, such as high volume, referral context, and specific staffing patterns, may not transfer directly to lower-level facilities, private-sector settings, or rural hospitals. This transferability concern was also encountered by Oluka [23], in his study conducted at a national tertiary referral hospital (Mulago). Several additional sampling and design features may have shaped these findings’ generalisability. The two-stage recruitment process, in which women were approached only at a scheduled review appointment following discharge, may have systematically under-sampled women most fearful of re-engaging with the health system plausibly those with induced rather than spontaneous abortion presentations, who in Uganda’s restrictive legal context are least likely to attend follow-up. This would tend to concentrate the sample toward more socially acceptable presentations, potentially understating the most severe stigma-related experiences the study aimed to capture. Relatedly, the 33% refusal rate among approached women (eight of twenty-four) is not further characterised in this manuscript; if declination was non-random and concentrated among women experiencing greater shame, fear of legal exposure, or more traumatic care, the reported themes may under-represent the most distressing EPAC experiences. The interview setting itself conducted on hospital premises during a scheduled clinical review appointment, while participants remained dependent on the facility for ongoing recovery, may have introduced institutional social-desirability pressure distinct from general social desirability bias; the presence of multiple negative accounts across the dataset demonstrates that total suppression of criticism did not occur, but cannot establish that the intensity or frequency of provider-directed criticism was unmoderated, and the prominence of gratitude-framed narratives in Theme 1 and reassurance-framed narratives in Theme 3 should be interpreted with this in mind. A further, distinct power differential existed between the research team and participants: beyond the clinical-institutional insider position discussed in Researcher positionality above, the wider research team was academically and institutionally senior to a participant group of whom 62.5% had at most lower secondary education (Table 1), which may have differentially suppressed criticism of individual providers (Theme 4) relative to criticism of abstract systems (Theme 5). The 14-day post-discharge recruitment threshold may still have been too early to capture the full psychological consequences of fertility uncertainty and emotional processing of guilt or grief, which tend to become more salient over longer periods; because recruitment occurred only at or after this point, all participants had already reached, or nearly reached, the standard 1–2 week post-abortion follow-up appointment by the time of interview, so accounts describing an absence of follow-up contact (Theme 6) reflect experience up to that point rather than a longer-term absence of follow-up. Finally, data were collected across a single two-month period at one facility; conditions prevailing during that period, such as stock availability, staffing, and patient flow, may not represent the full range of conditions over time. Longitudinal follow-up would provide a more complete picture of recovery trajectories and lasting dimensions of the care experience. Several further characteristics were not captured as sampling variables or systematically reported. Geographic origin relative to KNRH was not recorded; given KNRH’s role as a national referral centre serving both peri-urban Kampala and more distant catchment areas, referral pathway experiences (Theme 5) may differ systematically for women travelling from outside greater Kampala in ways not represented in the current sample. Religious affiliation was similarly not recorded; as Christian and Islamic moral frameworks are known to shape guilt, self-blame, and interpretation of judgmental clinical communication in Uganda, the moral framing evident in Themes 2 and 4 may reflect the dominant religious composition of this sample rather than the full range of EPAC experience. Relatedly, whether each participant presented following spontaneous pregnancy loss or induced abortion was not systematically recorded (see Study population, above); because these presentations can carry different emotional, social, and legal meanings even where clinical management converges, the guilt, stigma, and moral-vulnerability findings in Themes 2 and 4 should be read as grounded in the subset of narratives where this context was disclosed, rather than as characterising the sample as a whole. Uterine evacuation procedure type (e.g., manual vacuum aspiration, electric vacuum aspiration, sharp curettage, or misoprostol-only management) was not systematically recorded, limiting the clinical precision with which the procedural pain findings in Theme 2 can be linked to specific management approaches. Finally, translation and back-translation of Luganda-language interviews were conducted by the interviewers themselves rather than by translators independent of the research team, which is noted here as a methodological limitation. Relatedly, although the study protocol included a defined procedure for referring participants for clinical assessment when concerning symptoms were disclosed mid-interview, no case during data collection required this procedure to be invoked (see Theme 6, above). Two further points concern the pathway of care within the study site itself. First, the interview guide contained no dedicated question on referral or transfer at any stage; pre-arrival referral content emerged through the open-ended opening question and its “step by step” probe, while referral or transfer between departments within KNRH once admitted (e.g., reception, the gynaecological emergency unit, and theatre) was neither asked about nor volunteered. The findings reported under Theme 5 should therefore be read as reflecting what participants spontaneously narrated about the pre-arrival pathway, not as a systematic or design-intended account of referral experience, and the absence of intra-facility content should not be read as evidence that no such experiences occurred. Future research on EPAC at KNRH could add a dedicated question on intra-facility transfer specifically. Second, participant accounts distinguish providers descriptively (for example, as “the doctor” or “a female doctor”) but provider cadre (e.g., consultant, resident, intern, midwife, or nursing officer) was not systematically recorded during data collection, so this analysis cannot establish whether experiences of pain management, communication, or dignity differed systematically by provider cadre, as opposed to by individual provider or by moment in the care pathway.

## Conclusions

Among the sixteen women interviewed, EPAC at KNRH was experienced not as a single clinical event but as a multi-dimensional encounter shaped by bodily vulnerability, provider behaviour, structural conditions, and the social context of abortion. Within this sample, survival was necessary but not sufficient as a measure of quality. Communication, pain management, and privacy were core quality dimensions, not supplementary comforts. Structural barriers, including delays, shortages, out-of-pocket costs, and fragmented referrals, constrained dignified care. The experience of EPAC extended beyond discharge, with recovery experienced by these participants as largely unstructured, with fertility uncertainty persisting without follow-up.

Improving the quality of EPAC requires a patient-centred approach that integrates clinical effectiveness with respectful, non-judgmental communication, systematic pain management, improved triage, better use of privacy measures, reliable availability of essential medicines and blood products, stronger referral coordination, and structured discharge counselling and follow-up. Provider sensitisation should address stigma, which this study found to shape how care was communicated and experienced, though its causal contribution to objective care quality was not directly tested. Future research should examine EPAC experiences across the full referral pathway, include provider perspectives, and assess long-term physical, psychological, and reproductive outcomes after emergency care.

## Supporting information

CONSOLIDATED CRITERIA FOR REPORTING QUALITATIVE RESEARCH (COREQ)

: Interview guide: for women who have received health care for management of abortion related complications at KNRH-English version

## Author contributions

Conceptualisation: KSS, HK, KA

Data curation: KSS, AGJ, BA, IP, KB

Formal analysis: KSS

Investigation: KSS, AGJ, BA, IP, KB

Methodology: KSS, HK, KA

Project administration: KSS, AGJ, BA, IP, KB

Resources: KSS, AGJ, BA, IP, KB

Supervision: HK, KA

Validation: HK, KA

Visualisation: KSS

Writing – original draft preparation: KSS

Writing – review and editing: KSS, HK, KA

## Data availability statement

All data underlying this study consist of in-depth interview transcripts containing sensitive personal health information shared by participants under assurances of confidentiality. Full transcripts cannot be shared publicly, as doing so would violate participant consent agreements and risk identification of individuals who disclosed stigmatised health experiences. Anonymised thematic data are available on reasonable request from Makerere University College of Health Sciences (Department of Obstetrics and Gynaecology), P.O. Box 7072, Kampala, Uganda, subject to SOMREC approval (reference Mak-SOMREC-2025-913) and compliance with applicable data protection requirements. Requests should reference approval number Mak-SOMREC-2025-913. The analytical documentation underlying the reported themes, including the coding log and analytic memo trail maintained throughout analysis, is available from SOMREC under the same conditions and approval reference.

## Acknowledgments

The authors thank the women who shared their experiences despite the emotional and physical difficulties they had undergone. Their voices made this work possible. We acknowledge Kawempe National Referral Hospital for providing the research setting. We thank the research assistants who supported data collection and the Department of Obstetrics and Gynaecology, Makerere University, for academic mentorship throughout this work.

## Supporting information

S1 Interview guide: for women who have received health care for management of abortion related complications at KNRH-English version

S2 COREQ Checklist. COREQ (Consolidated Criteria for Reporting Qualitative Research) checklist for the study.

