## Supplementary material for "Women’s experiences of emergency post-abortion care at Kawempe National Referral Hospital, Uganda: a qualitative descriptive study": CONSOLIDATED CRITERIA FOR REPORTING QUALITATIVE RESEARCH (COREQ)

**S2 File_CONSOLIDATED CRITERIA FOR REPORTING QUALITATIVE RESEARCH (COREQ)**

**Target journal:** PLOS ONE | Ethics: SOMREC, Mak-SOMREC-2025-913

| **No.** | **Item** | **Guide question / description** | **Response** | **Reported on Page(s)** |
| --- | --- | --- | --- | --- |
| **DOMAIN 1: Research team and reflexivity** | | | | |
| ***Personal characteristics*** | | | | |
| **1** | Interviewer / facilitator | Which author/s conducted the interview or focus group? | In-depth interviews were conducted by the principal investigator (Kawungu Saad Sessimba, MBChB) supported by trained research assistants who facilitated participant approach, consent, and data collection. | Pages 7, 12 |
| **2** | Credentials | What were the researcher's credentials, e.g. PhD, MD? | Principal investigator: MBChB (Makerere University), postgraduate candidate in MMed Obstetrics and Gynecology. Supervisors (Herbert Kayiga: MBChB, MMed, MPH, PhD; Keesiga Annette: MBChB, MMed, MPH) stated on cover page and title page. | Not reported |
| **3** | Occupation | What was their occupation at the time of the study? | Principal investigator: postgraduate student, Department of Obstetrics and Gynecology, CHS, Makerere University. Research assistants: registered midwives at KNRH. | Page 1 |
| **4** | Gender | Was the researcher male or female? | Not explicitly stated in the manuscript. The researcher is however male. | Not reported |
| **5** | Experience and training | What methodological experience and training did the researcher have? | The principal investigator is enrolled in a postgraduate research degree. Research assistants underwent standardized training in study protocol, informed consent, and semi-structured interviewing. Formal methodological qualifications not detailed beyond the degree program. | Pages 7, 12 |
| ***Relationship with participants*** | | | | |
| **6** | Relationship established | Was a relationship established prior to study commencement? | No prior relationship. Two-stage approach: women were identified at discharge (Stage 1); the research team approached them at the scheduled follow-up review visit (Stage 2). First contact for study purposes occurred during the discharge stage. | Pages 7, 12 |
| **7** | Participant knowledge of the interviewer | What did participants know about the researcher e.g. personal goals, reasons for doing the research? | Before consent was obtained, each participant was given a verbal explanation of the study purpose, the researcher's role as a postgraduate student, and the voluntary nature of participation with no effect on clinical care. | Page 7 |
| **8** | Interviewer characteristics | What characteristics were reported about the interviewer/facilitator? | Research assistants were trained registered midwives familiar with KNRH's clinical environment and able to conduct interviews in English and Luganda. Interviews were conducted individually in a private consultation room. | Pages 7, 12 |
| **DOMAIN 2: Study design** | | | | |
| ***Theoretical framework*** | | | | |
| **9** | Methodological orientation and theory | What methodological orientation was stated to underpin the study, e.g. grounded theory, discourse analysis, ethnography, and phenomenology? | Qualitative descriptive design, without commitment to a specific phenomenological, grounded-theory, or narrative analytic tradition. Inductive thematic analysis was used for coding and theme generation. The Socio-Ecological Model (SEM) was applied as an organising interpretive framework at later stages of analysis to situate findings across individual, interpersonal, facility, and community levels. | Pages 7, 13 |
| ***Participant selection*** | | | | |
| **10** | Sampling | How were participants selected, e.g. purposive, convenience, consecutive, snowball? | Purposive sampling with maximal variation strategy, ensuring diversity in age, marital status, educational attainment, occupation, and parity to maximize analytical breadth. | Page 11 |
| **11** | Method of approach | How were the participants approached? E.g. face to face, telephone, mail | Two-stage face-to-face approach: (1) at discharge from KNRH, eligible women were identified and a review appointment confirmed; (2) at the review visit, the research assistant approached participants face to face, provided a verbal briefing, and obtained written informed consent. | Page 12 |
| **12** | Sample size | How many participants were in the study? | Sixteen (16) participants. Sample size determined by data saturation, assessed narratively, not by a priori power calculation or by phenomenological criteria such as depth of individual accounts. | Pages 11, 13 |
| **13** | Non-participation | How many people refused to participate or dropped out? Reasons? | Eight women declined participation during March–April 2026, citing time constraints or personal concerns. No participants withdrew after enrolment. No dropouts occurred. | Page 12 |
| ***Setting*** | | | | |
| **14** | Setting of data collection | Where was the data collected? E.g. home, clinic, workplace | A private consultation room within the gynecological emergency unit, KNRH, Kampala, Uganda. Setting was chosen to ensure privacy and minimize disruption to clinical care. | Pages 9, 12 |
| **15** | Presence of non-participants | Was anyone else present besides the participants and researchers? | Not explicitly reported. Interviews were conducted individually in a private room, indicating no other patients or staff were present during data collection. | Page 12 |
| **16** | Description of sample | What are the important characteristics of the sample, e.g. demographic data, date? | Participants aged 18–45 years. Full socio-demographic characteristics (age, marital status, education, occupation, number of children) presented in Table 1. Data collected March–April 2026. | Pages 10, 16 |
| ***Data collection*** | | | | |
| **17** | Interview guide | Were questions, prompts, guides provided by the authors? Was it pilot tested? | Semi-structured interview guide used, opening with an account of the participant's overall experience of care, including probes on privacy, confidentiality, and feeling respected, and continuing with: prior knowledge and expectations of care; interactions with health workers; communication and information-sharing; support from family and others; community attitudes and stigma; recommendations for the health system and policy; and a final open reflection. Guide provided as S1 File. Pilot testing was done. | Page 12 |
| **18** | Repeat interviews | Were repeat interviews carried out? If yes, how many? | Not applicable. Each participant was interviewed once. | N/A |
| **19** | Audio/visual recording | Did the research use audio or visual recording to collect the data? | Yes. Interviews were audio-recorded with participant consent and transcribed verbatim. Back-translation procedures were applied where interviews were conducted in Luganda. | Page 12 |
| **20** | Field notes | Were field notes made during and/or after the interview? | Not reported in the manuscript. | N/A |
| **21** | Duration | What was the duration of the interviews or focus group? | Duration of individual interviews was reported. Data collection took place across a two-month period (March–April 2026). | Page 12 |
| **22** | Data saturation | Was data saturation discussed? | Yes. Sampling was guided by the principle of data saturation; collection continued until no new codes, categories, or themes emerged from additional transcripts. | Page 11 |
| **23** | Returning transcripts | Were transcripts returned to participants for comment and/or correction (member checking)? | Not reported. | N/A |
| **DOMAIN 3: Data analysis and findings** | | | | |
| ***Data analysis*** | | | | |
| **24** | Number of data coders | How many data coders were used? | Not explicitly stated. But 67 codes were generated with 20 sub themes identified  And 6 final themes generated | Page 13; Table 2 on page 16 |
| **25** | Description of the coding tree | Did authors provide a description of the coding tree? | Yes. Table 2 (p. 16) presents all six themes, their sub-themes, and corresponding meaningful units (codes), forming a complete coding structure. | Page 16; Table 2 on page 16 |
| **26** | Derivation of themes | Were themes identified in advance or derived from the data? | Themes were derived inductively. Thematic analysis moved iteratively between participants' language and broader interpretive concepts. The SEM was applied subsequently as an organising framework rather than a deductive coding template. | Page 13 |
| **27** | Software | What software, if applicable, was used to manage the data? | No qualitative data management software is reported. Analysis was conducted through direct iterative engagement with transcripts. | Page 13 |
| **28** | Participant checking | Did participants provide feedback on the findings? | Not reported. | N/A |
| ***Reporting*** | | | | |
| **29** | Quotations presented | Were participant quotations presented to illustrate the themes/findings? | Yes. Verbatim quotations presented throughout the Results section for all six themes and their sub-themes, in italics, attributed by participant code (e.g., IDI_16). | Pages 18–25 |
| **30** | Data and findings consistent | Was there consistency between the data presented and the findings? | Yes. Each theme and sub-theme is directly supported by verbatim quotations. Table 2 maps codes to sub-themes and themes. Divergent accounts (e.g., co-existence of positive survival evaluation with reports of pain, privacy violations, and financial burden) are explicitly acknowledged. | Pages 16–25 |
| **31** | Clarity of major themes | Were major themes clearly presented in the findings? | Yes. Six clearly labelled major themes, each with a dedicated Results sub-section, descriptive narrative, and supporting quotations. Table 2 provides a full structured overview of all themes, sub-themes, and codes. | Pages 16–25; Table 2 on page 16 |
| **32** | Clarity of minor themes | Is there a description of diverse cases or discussion of minor themes? | Yes. Divergent experiences are foregrounded throughout Results and Discussion — including contrasting accounts of positive and negative communication, presence vs. absence of social support, and post-discharge uncertainty. Minor themes such as self-blame, reproductive anxiety, and selective social disclosure are discussed. | Pages 18–29 |

Note N/A- not applicable for study design
