## Supplementary material for "Women’s experiences of emergency post-abortion care at Kawempe National Referral Hospital, Uganda: a qualitative descriptive study": : Interview guide: for women who have received health care for management of abortion related complications at KNRH-English version

**S1 File:** Interview guide: for women who have received health care for management of abortion related complications at KNRH-English version

**STUDY TITLE**: Women’s Experiences of Emergency Post-Abortion Care at Kawempe National Referral Hospital, Uganda.

**Interviewee Characteristics:**

Initials:

Age:

Marital status:

Education-level:

Occupations:

Parity:

1. Opening Question: Tell me about the emergency post-abortion care you received at Kawempe National Referral Hospital. How would you describe your experience, from the moment you realised something was wrong until you left the hospital?

Probes:

•	What happened, step by step?

•	What stands out most to you about that time, and why

•	Did you feel you had privacy? Beyond physical privacy, did you feel confident that your personal information and the reason for your visit would be kept confidential? Did you feel safe from judgment or harm while you were at the hospital? How did that affect your experience?

•	What did staff say or do that made you feel respected or not respected? Can you describe a specific moment and how that affected your experience?

2. Prior Knowledge and Expectations: Before this experience, what did you know or expect about care for complications like yours?

Probes:

•	What had you heard from others - friends, family, or community members about this kind of care?

•	Had you heard any stories—positive or negative—about Kawempe hospital specifically?

•	Describe how that knowledge or lack of it impacted your experience of care, including your feelings about coming to the hospital.

3. Interactions with Health Workers: Tell me about your interactions with the health workers – the nurses, doctors, and others who cared for you.

Probes:

•	Can you explain things in a way you could understand? How did that- or lack of it impact your experience?

•	Did you feel involved in decisions about your care? Describe how that involvement—or lack of it—made you feel, and whether it relates to your everyday life or sense of dignity. Can you describe a specific moment?

4. Communication and Information: Describe how you came to understand what was happening to you and what treatment you needed.

Probes:

•	Was there anything you wanted to ask and didn’t ask? Describe what happened in that moment—what stopped you, and how did that impact your experience?

•	Did you feel you could ask questions if you wanted to? Why or why not?

5. Support from Family and Others: Who was with you – or who did you wish was with you – during this time?

Probes:

•	Did you receive emotional support from a partner, health workers or others? How did that support – or lack of it – affect you during and after your time in the hospital?

•	Did you tell anyone you were coming to the hospital? Why or why not, and how did that decision affect your experience?

6. Community Attitudes and Stigma: In your community, how are women viewed when they seek care like this – and did those views affect you personally? If so, how? If not, why do you think that is?

Probes:

•	Did you feel shame, judgment, or guilt at any point during this experience? Where did those feelings come from—inside yourself, from others, or from the environment around you?

•	How did those feelings affect your decisions, your interactions with staff, or your overall experience?

7. Health System and Policy Context: Based on your experience, what do you think should change about how emergency care is provided to women with pregnancy complications in Uganda?

Probes:

•	If you were asked to advise the Ministry of Health or Kawempe Hospital, what would you want them to know or change?

8. Final Reflection: Is there anything else about your experience that you feel is important for me to understand – anything we haven't touched on that you want to share?

Probe

•	If you could change just one thing about the experience you had, what would it be and why? What would that new experience look like for the next woman who comes here?

•	Was there anything that went well—anything you think they should definitely keep doing? Why was that important to you?
